# Mid-term subclinical myocardial injury detection in patients recovered from COVID-19 according pulmonary lesion severity

**DOI:** 10.1101/2022.03.14.22272130

**Authors:** Ikram Chamtouri, Rania Kaddoussi, Hela Abroug, Mariem Hafsa, Mabrouk Abdeleli, Nessrine Fahem, Taha Lassoued, Asma Ben Abdallah, Asma Achour, Asma Migaou, Sawssen Cheikhm’hamed, Walid Jomaa, Asma Sriha, Khaldoun Ben Hamda, Faouzi Maatouk

**Affiliations:** Cardiology B department, Fattouma Bourguiba University Hospital, Monastir, Tunisia; Pneumolgy department, Fattouma Bourguiba University Hospital, Monastir, Tunisia; Preventive department, Fattouma Bourguiba University Hospital, Monastir, Tunisia; Pneumolgy department, Ksar Helal hospital and Moknine Hospital; Radiology department, Fattouma Bourguiba University Hospital, Monastir, Tunisia

## Abstract

**Background:** Severe acute respiratory syndrome coronavirus 2 (SARS-CoV 2) may cause damage of the cardiovascular system during the acute phase of infection. However, Recent studies described a mid and long-term subtle cardiac injuries after recovery from acute Coronavirus disease 19 (COVID-19).The aim of this study was to determine the relationship between the severity of chest computed tomography (CT) lesions and the persistence of subtle myocardial injuries at mid-term follow-up of patients recovered from COVID-19 infection.

**Methods:** All COVID-19 patients were enrolled prospectively in this study. Sensitive troponin T (hsTnT) and chest CT scan was performed in all patients at the acute phase of Covid-19 infection. At the mid-term follow up, conventional transthoracic echocardiograph and global longitudinal strain (GLS) of left and right ventricles (LV and RV) were determined and compared between patients with chest CT scan lesions less than 50% (Group 1) and those with severe chest CT scan greater or equal to 50% (Group 2).

**Results:** The mean age was 55 more or less than 14 years. Both LV GLS and RV GLS were significantly decreased in the group 2 (p=0.013 and p=0.011, respectively). LV GLS value more than -18% was noted in 43% of all the patients and RV GLS value more than -20% was observed in 48% of them. The group with severe chest CT scan lesions included more patients with reduced LV GLS and reduced RV GLS than the group with mild chest CT scan lesions (G1:29% vs. G2:57%, p=0.002) and (G1:36% vs. G2:60 %, p=0.009) respectively).

**Conclusion:** Patients with severe chest CT scan lesions are more likely to develop subclinical myocardial damage. TTE could be recommended in patients recovering from COVID-19 to detect subtle LV and RV lesions.

**Trial registration:** The cohort of patients is a part of the research protocol (IORG 00093738 N°102/OMB 0990-0279) approved by the Hospital Ethics Committee.

## Introduction

Coronavirus Disease2019 (COVID-19) is an emergent infection caused by the new coronavirus SARS-COV 2 [1]. Since its appearance, it has caused millions of deaths by affecting several organs. Although the respiratory system is the most affected, cardiac involvement seems to have a big role in prognosis [2]. Performing transthoracic echocardiography (TTE), especially in critical cases may help to evaluate prognosis and guide therapeutic management. Many studies [4-5] have shown the importance of longitudinal strain study in hospitalized patients with confirmed COVID-19 infection. However, only few ones have studied its contribution in the detection of infra-clinical myocardial injuries after the recovery from acute infection. Myocardial speckle tracking study by measuring Left ventricle global longitudinal strain (LV GLS) and right ventricle global longitudinal strain (RV GLS) could be helpful in the determination of subclinical myocardial damage [3]. The aim of this study was to reveal cardiac abnormalities at mid-term follow-up in patients recovered from COVID-19 using LV GLS and RV GLS and to compare the degree of LV GLS and RV GLS alteration between patients having already severe chest computed tomography (CT) scan lesions ≥ 50% and those with chest CT scan lesions <50% at the acute phase of infection.

## Methods

### Study design and population

This is a cohort multicentric study conducted in three COVID units and including consecutive North Africans patients hospitalized for COVID-19 infection and aged > 18 years. COVID-19 diagnosis was based on the presence of a real-time reverse-transcription polymerase chain reaction (RT-PCR). All the patients had clinical examination, blood tests including sensitivity troponin T (hsTnT), C-reactive protein, D-dimeres, hematocrit, serum creatinine, N-Terminal prohormone of Brain Natriuretic Peptide (NT-proBNP) and chest CT scan. Patients were divided into two groups. Group 1: those with Chest CT scan lesions less than 50% and Group 2: those with chest CT scan lesions equal to or greater than 50%. Three months after recovery, TTE with longitudinal strain study was performed. Non-inclusion criteria were pulmonary embolism and a hsTnT serum level> the 99^th^ percentile (0.014 ng/mL) [4] during the acute phase of COVID-19 infection. A level of hs-cTnT level more than the 99^th^ percentile (0.014 ng/mL) at three months, poor acoustic windows, severe valvular heart disease, LVEF <50%, and arrhythmia represented exclusion criteria (figure 1).

**Figure 1:**
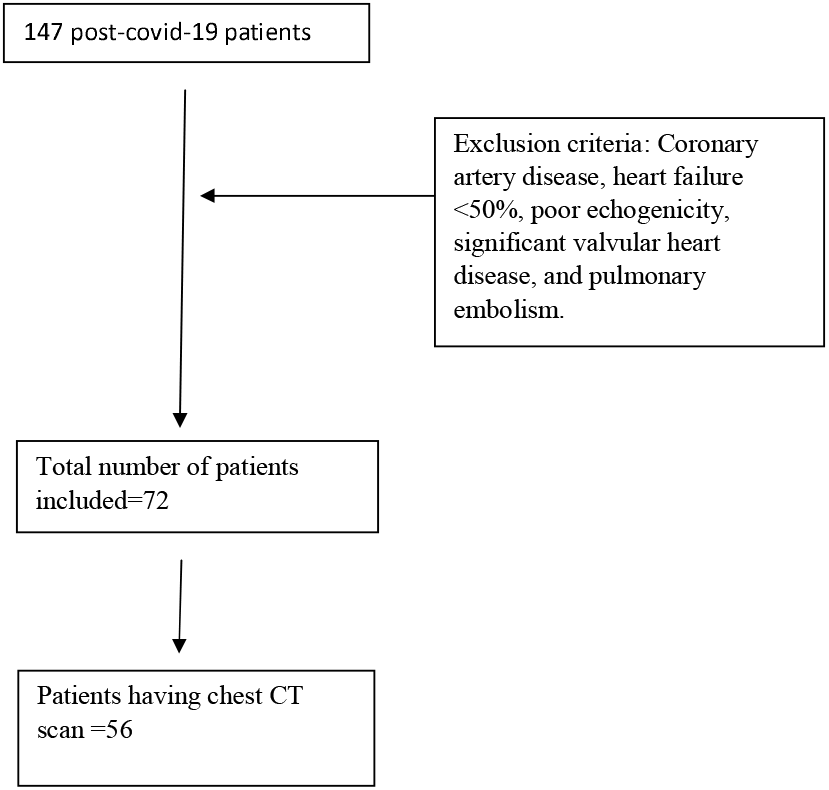
Study population

All participants had signed an informed consent.

### Data sources and measurement

Demographic, biological and scanner data were collected during hospitalization in the COVID-19 units.

At three months of follow-up, hsTnT and TTE were performed in all participants. TTE using the Vivid E9 General Electric Medical System was performed by the same operator. Conventional TTE parameters, including LV end-systolic and LV end-diastolic diameters (LVESD and LVEDD), interventricular septum (IVS), and LV ejection fraction (LVEF) were measured.

Diastolic LV function was evaluated. RV systolic function was performed by using peak right ventricular systolic myocardial velocity (S wave) and tricuspid annular plane systolic excursion (TAPSE). Atria areas and systolic pulmonary blood pressure(SPBP) were measured. Offline speckle tracking analyses were performed in all participants using Echopac software version 112 and based on apical two, three, and four chambers views for LV study. GLS was obtained after tracking the endocardial borders and by calculating the mean peak systolic strain values of the 17 segments. LV GLS and bull’s eye plot were obtained (figure 2). A value of LV GLS more than -18% was regarded as pathological [5]. RV GLS was obtained from the apical four-chamber view by tracking the RV free wall endocardial borders. RV GLS was calculated from the mean strain values of the RV three segments (basal, mid, and apical) (figure 3). RV GLS more than -20% was considered as pathological [6]

**Figure 2:**
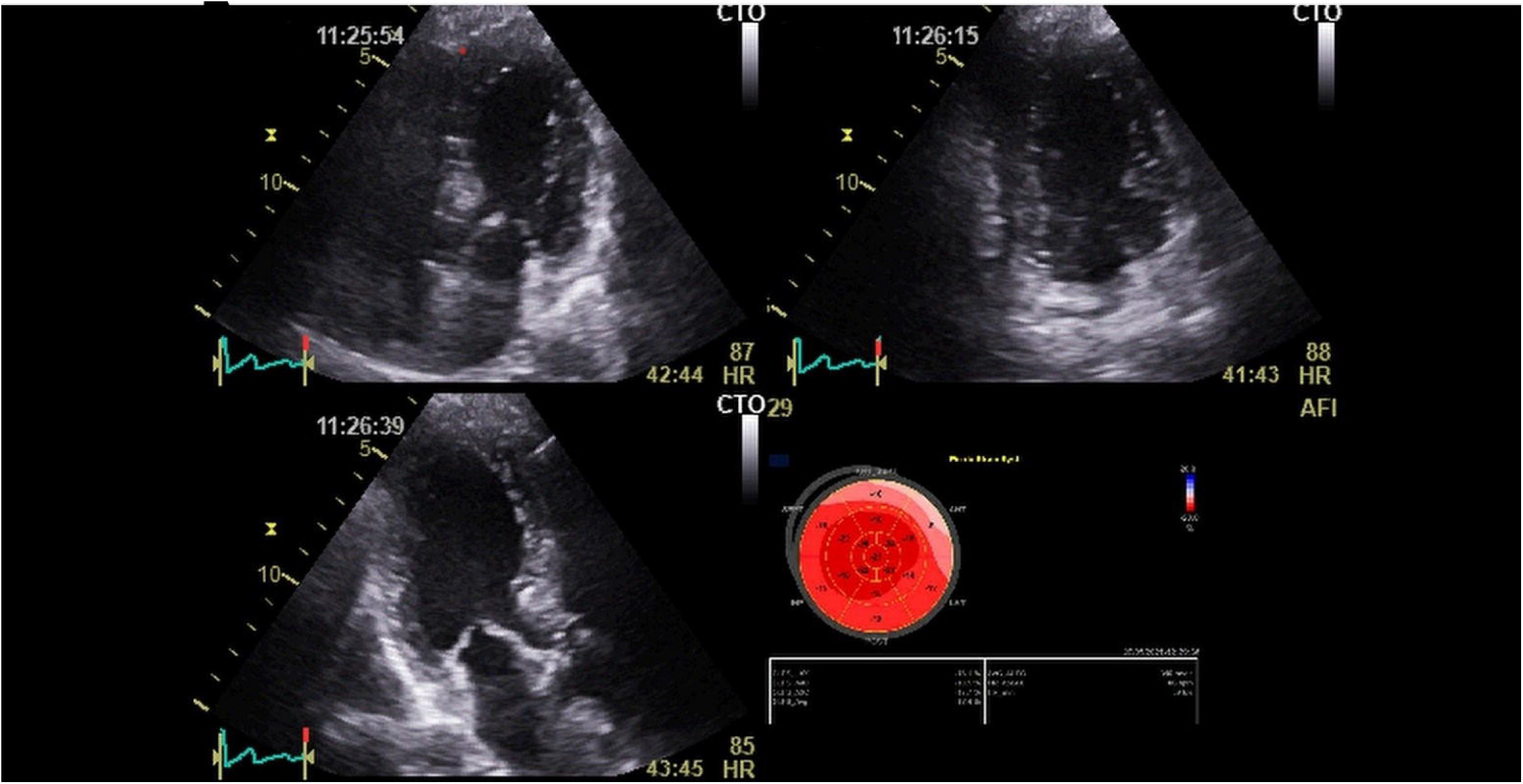
LV GLS and bull’s eye plot recovered patient from COVID-19 infection.

**Figure 3:**
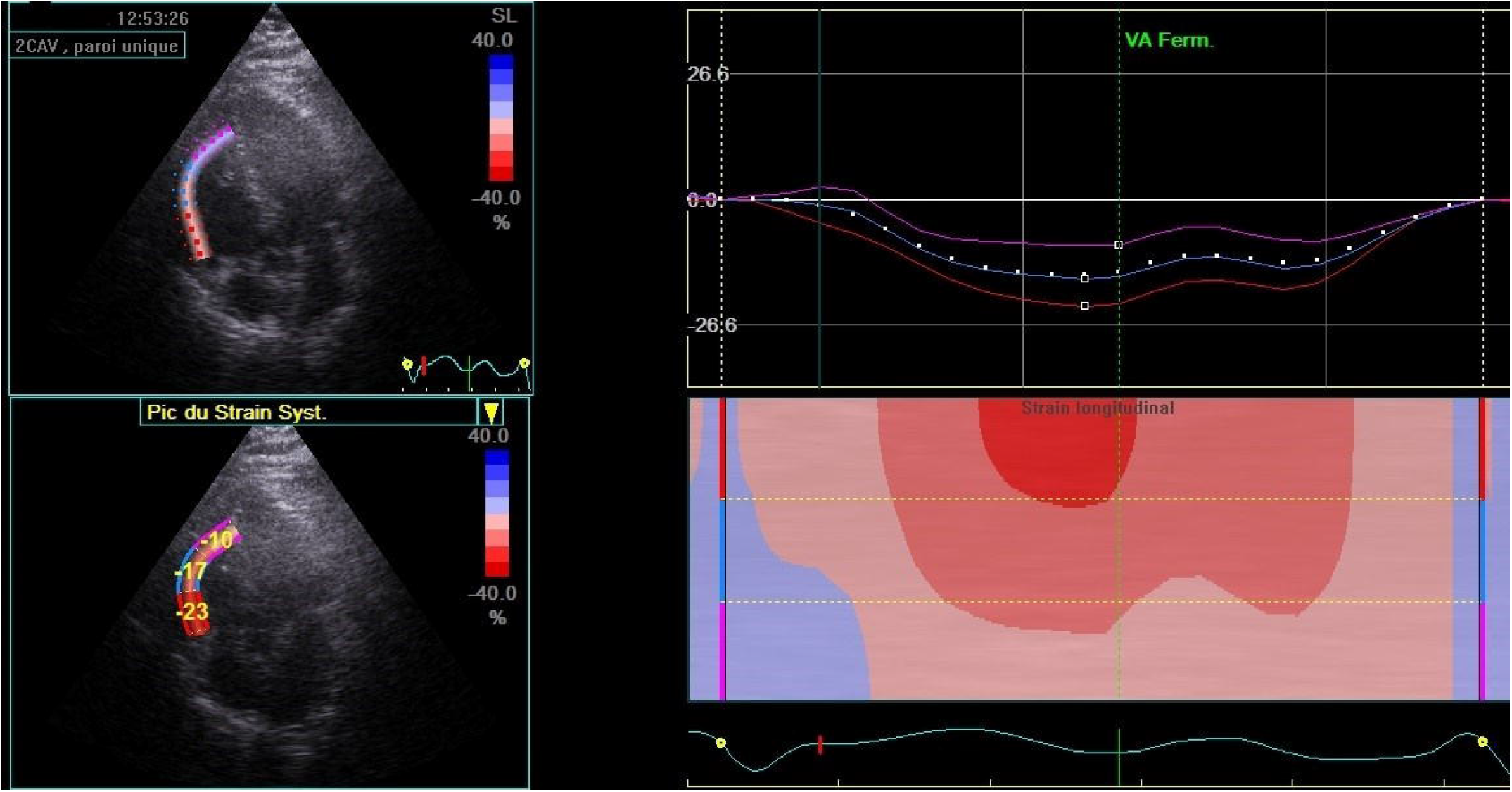
RV GLS calculated from the mean strain values of the RV three segments (basal, mid, and apical) in patient recovered from COVID-19 infection.

### Statistical analysis

Statistical analyses were performed using SPSS (Statistical Program for Social Sciences) version 20.0. Kolmogorov–Smirnov test was used to confirm the normal distribution of continuous variables. Categorical variables were presented as frequency rates and percentages. The Chi-Square test (χ2 test) or the Fisher exact test were used to compare frequencies. The normally distributed data were expressed as mean ±standard deviation (SD) and the student t test was performed to compare means in two independent samples. The non-normally distributed data were presented as median with interquartile range (IQR) and compared by the Mann-Whitney U test. Statistical tests were two-tailed, and a P value of <0.05 was considered statistically significant.

## Results

A total of 147 patients were initially enrolled, among them 37 had exclusion criteria. Thus, a total of 111 patients were included in the final analysis. Group 1 (55 patients) with mild chest CT scan lesions (<50%) and group 2 (56 patients) with severe chest CT scan ≥ 50% (Figure 1). Baseline data of the overall population and the two groups are presented in Table 1. No significant differences were noted between the two groups in all baseline characteristics. Compared to group 1, a higher prevalence of ground glass opacity was noted in group 2 (86. 2% vs. G2: 94.4%; p= 0.05).

**Table 1:**
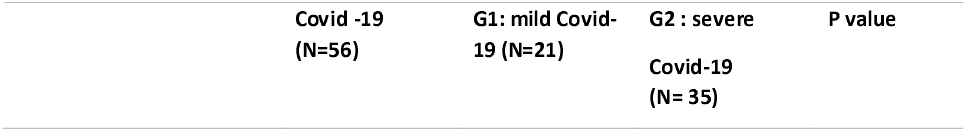
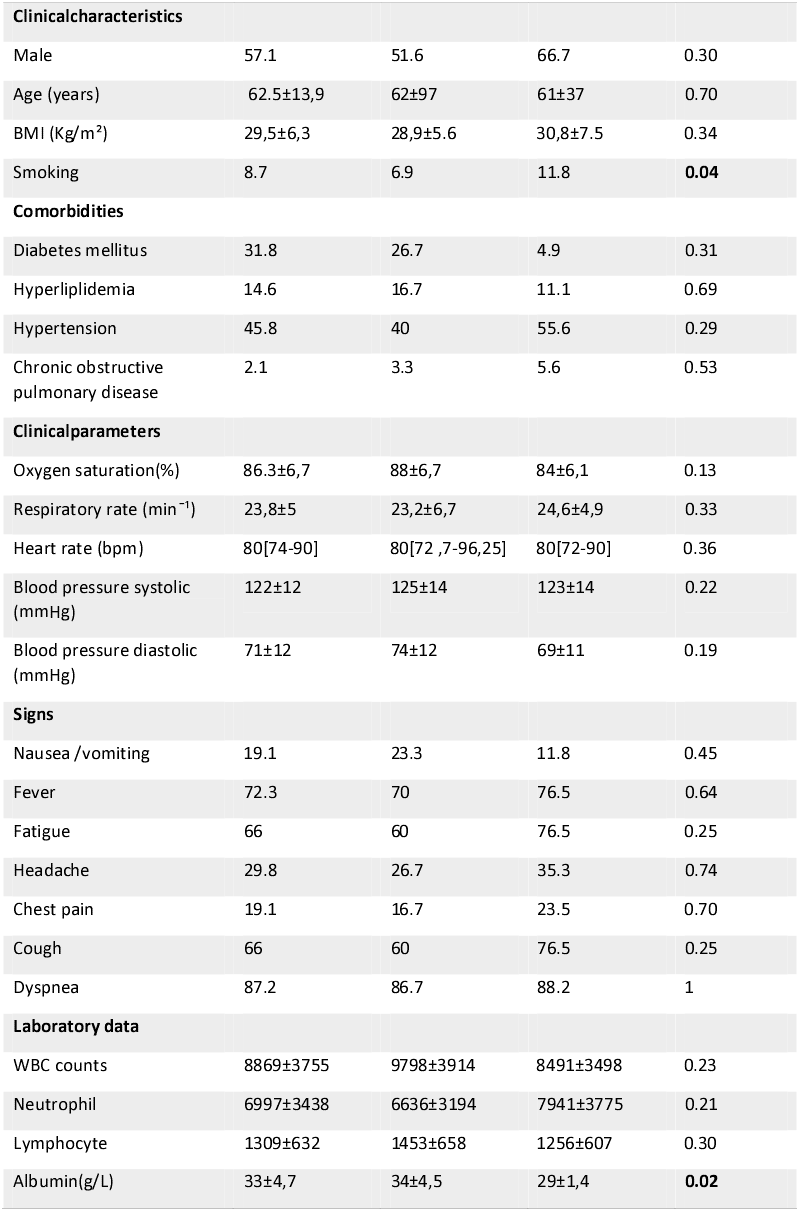
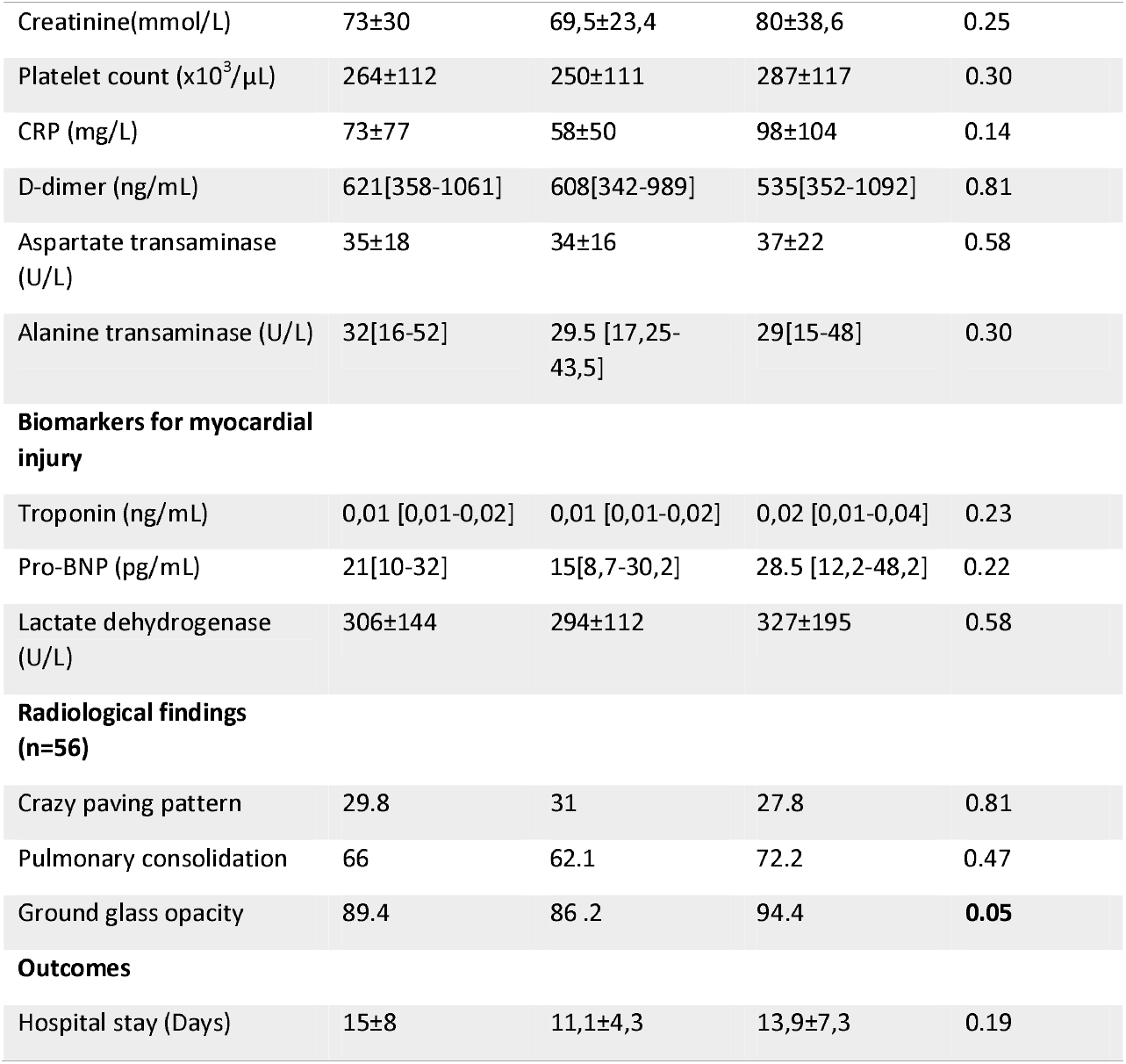
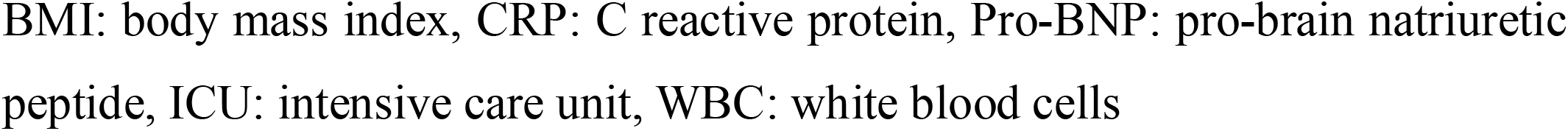
Patients’ characteristics, symptoms, comorbidities, and laboratory findings.

|  | Covid -19<br>(N=56) | G1: mild Covid-19 (N=21) | G2 : severe<br>Covid-19<br>(N= 35) | P value |
| --- | --- | --- | --- | --- |

| Clinical characteristics |  |  |  |  |
| --- | --- | --- | --- | --- |
| Male | 57.1 | 51.6 | 66.7 | 0.30 |
| Age (years) | 62.5±13,9 | 62±97 | 61±37 | 0.70 |
| BMI (Kg/m<sup>2</sup>) | 29,5±6,3 | 28,9±5.6 | 30,8±7.5 | 0.34 |
| Smoking | 8.7 | 6.9 | 11.8 | **0.04** |
| Comorbidities |  |  |  |  |
| Diabetes mellitus | 31.8 | 26.7 | 4.9 | 0.31 |
| Hyperlipidemia | 14.6 | 16.7 | 11.1 | 0.69 |
| Hypertension | 45.8 | 40 | 55.6 | 0.29 |
| Chronic obstructive pulmonary disease | 2.1 | 3.3 | 5.6 | 0.53 |
| Clinical parameters |  |  |  |  |
| Oxygen saturation(%) | 86.3±6,7 | 88±6,7 | 84±6,1 | 0.13 |
| Respiratory rate (min<sup>-1</sup>) | 23,8±5 | 23,2±6,7 | 24,6±4,9 | 0.33 |
| Heart rate (bpm) | 80[74-90] | 80[72 ,7-96,25] | 80[72-90] | 0.36 |
| Blood pressure systolic (mmHg) | 122±12 | 125±14 | 123±14 | 0.22 |
| Blood pressure diastolic (mmHg) | 71±12 | 74±12 | 69±11 | 0.19 |
| Signs |  |  |  |  |
| Nausea /vomiting | 19.1 | 23.3 | 11.8 | 0.45 |
| Fever | 72.3 | 70 | 76.5 | 0.64 |
| Fatigue | 66 | 60 | 76.5 | 0.25 |
| Headache | 29.8 | 26.7 | 35.3 | 0.74 |
| Chest pain | 19.1 | 16.7 | 23.5 | 0.70 |
| Cough | 66 | 60 | 76.5 | 0.25 |
| Dyspnea | 87.2 | 86.7 | 88.2 | 1 |
| Laboratory data |  |  |  |  |
| WBC counts | 8869±3755 | 9798±3914 | 8491±3498 | 0.23 |
| Neutrophil | 6997±3438 | 6636±3194 | 7941±3775 | 0.21 |
| Lymphocyte | 1309±632 | 1453±658 | 1256±607 | 0.30 |
| Albumin(g/L) | 33±4,7 | 34±4,5 | 29±1,4 | **0.02** |
| Creatinine(mmol/L) | 73±30 | 69,5±23,4 | 80±38,6 | 0.25 |
| Platelet count (x10<sup>3</sup>/μL) | 264±112 | 250±111 | 287±117 | 0.30 |
| CRP (mg/L) | 73±77 | 58±50 | 98±104 | 0.14 |
| D-dimer (ng/mL) | 621[358-1061] | 608[342-989] | 535[352-1092] | 0.81 |
| Aspartate transaminase (U/L) | 35±18 | 34±16 | 37±22 | 0.58 |
| Alanine transaminase (U/L) | 32[16-52] | 29.5 [17,25-43,5] | 29[15-48] | 0.30 |
| **Biomarkers for myocardial injury** |  |  |  |  |
| Troponin (ng/mL) | 0,01 [0,01-0,02] | 0,01 [0,01-0,02] | 0,02 [0,01-0,04] | 0.23 |
| Pro-BNP (pg/mL) | 21[10-32] | 15[8,7-30,2] | 28.5 [12,2-48,2] | 0.22 |
| Lactate dehydrogenase (U/L) | 306±144 | 294±112 | 327±195 | 0.58 |
| **Radiological findings (n=56)** |  |  |  |  |
| Crazy paving pattern | 29.8 | 31 | 27.8 | 0.81 |
| Pulmonary consolidation | 66 | 62.1 | 72.2 | 0.47 |
| Ground glass opacity | 89.4 | 86 .2 | 94.4 | **0.05** |
| **Outcomes** |  |  |  |  |
| Hospital stay (Days) | 15±8 | 11,1±4,3 | 13,9±7,3 | 0.19 |

BMI: body mass index, CRP: C reactive protein, Pro-BNP: pro-brain natriuretic peptide, ICU: intensive care unit, WBC: white blood cells

All echocardiographic measurements are reported in table 2. LVEF was normal in the two groups. No significant differences were noted in conventional echocardiographic parameters such as LVEF, LV diameters and IVS between the two groups. No significant difference was revealed in term of LV diastolic parameters such as E/A and E/e’. There was no significant difference between the two groups regarding standard parameters of RV function and SPBP. Compared to group 1, both mean LV GLS and RV GLS were significantly decreased in group 2 (p=0.013 and p=0.011, respectively). Reduced LV GLS >-18% was noted in 43% of all patients and reduced RV GLS >-20% was observed in 48% of them. Group 2 included more patients with reduced LV GLS (57% vs. 29%; p=0.002) and reduced GLS RV (60 % vs. 36%, p=0.009) compared to group 1(Table 3).

**Table 2:**
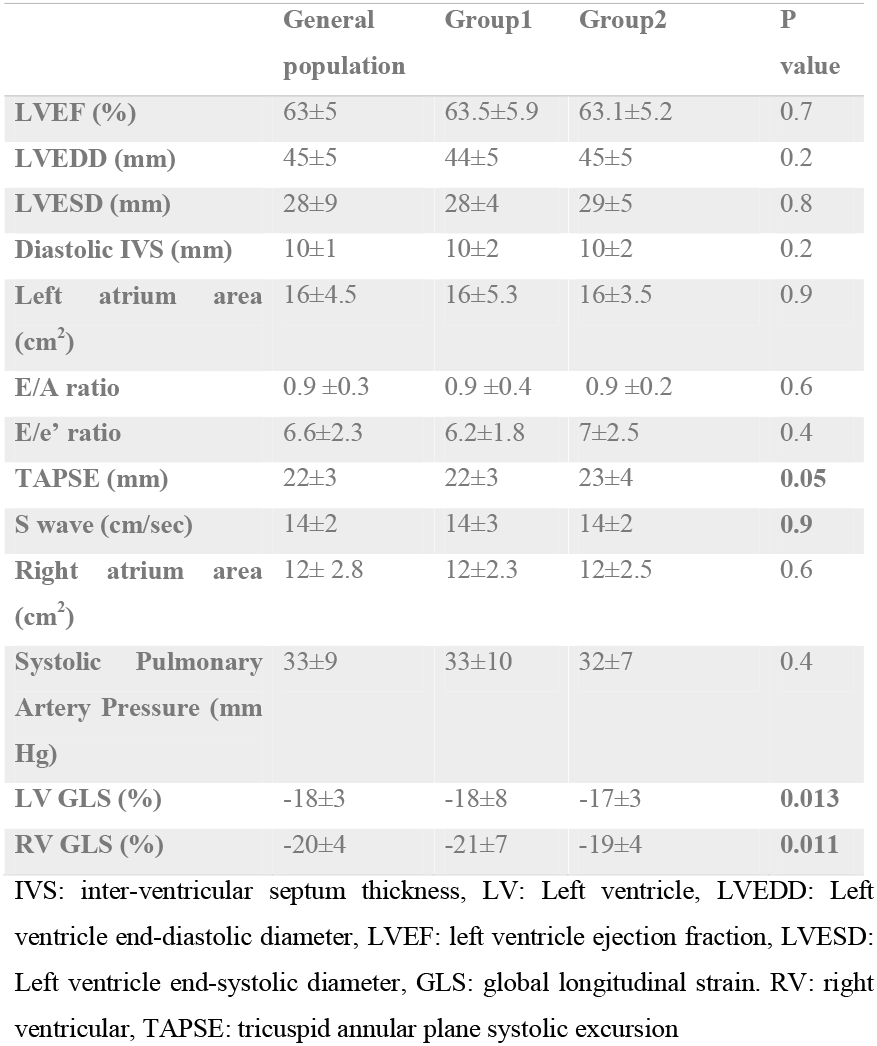
Echocardiography and myocardial strain measurement in post-Covid-19 patients. Comparison of conventional TTE parameters and GLS between the two groups.

**Table 3:** Comparison of LV GLS and RV GLS between the two groups.

|  | General<br>population | Group1 | Group2 | P value |
| --- | --- | --- | --- | --- |
| LV GLS >-18 (%) | 43% | 29% | 57% | 0.002 |
| RV GLS >-20 (%) | 48% | 36% | 60% | 0.009 |

GLS: global longitudinal strain, LV: left ventricular, RV: right ventricular,

## Discussion

Our study had tried to determine residual cardiac injury at mid-term follow-up among hospitalized patients recovered from COVID-19 and having no biological stigmata of myocardial injury, according the severity of previous pulmonary lesion. To the best of the authors’ knowledge, this is the first study evaluating the relation between biventricular strain in patients recovered from COVID-19 and lungs injury severity. According to a previous study, cardiovascular injuries due to COVID-19 infection could be manifested by acute coronary syndrome, myocarditis, arythmia, hypertension, pericarditis [2, 9-11]. Increased troponin level during acute phase of COVID-19 infection could define myocardial injury or other extra cardiac anomalies such as renal failure [12, 13]. Cardiac injury may be explained by the direct impact of the virus on myocardial tissue, hypercoagulation and thrombosis, endothelial dysfunction, and high cardiac stress due to hypoxemia and myocardial inflammation [3, 14].In this study elevated troponies level at the acute phase and at mid-term follow-up was an exclusion criterion.

Previous studies have shown that preserved LVEF in COVID-19 patients is associated with a decreased LV GLS, suggestive of occult myocardial injury, which is a marker of poor prognosis such as heart failure and mortality [15]. Despite eventual associated comorbidities like hypertension or diabetes, which may disrupt LV-GLS results, LV GLS has been demonstrated to be an independent predictor of bad outcomes in COVID-19 patients [16]. It could be reduced even in normotensive and non-diabetic patients [17, 18]. Previous studies have evaluated the correlation between LVGLS, COVID-19 symptoms, brain natriuretic protein (BNP), and troponin levels [19]. However, in the current study showed the impact of lungs injury severity on LV GLS alteration at three months after recovery from COVID-19 infection was investigated.

Reduced RV GLS has been demonstrated as a bad prognostic factor in patients with pulmonary embolism, pulmonary hypertension, and acute respiratory distress syndrome [20-22]. In COVID-19 patients, RV GLS alteration could be explained by RV inflammation and overload due to pulmonary embolism [23]. Several studies have reported that reduced RV GLS is associated with poor outcomes [8, 24]. In COVID-19 patients, RV free wall speckle tracking has a more excellent prognostic value than total RV strain (englobing inter-ventricular septum) which depends on LV systolic function and motion [7, 16, 25]. The current study showed a decrease in RV GLS compared to the reference value, with a significant reduction of RV GLS alteration according lungs injury severity. The potential hypothesis generated from this observation may be the concomitant factors inducing injury in both RV and lungs including the higher inflammatory burden, hypoxemia, and ventilation-induced injury, leading to subclinical RV injury out of the pulmonary embolism context [3, 26].

### Study limitations

Our study has some limitations. First, three dimension echocardiography as well as circonferencial and radial strain were not performed. Second, echocardiography was not performed during acute phase of COVID-19 infection. Moreover, cardiac magnetic resonance imaging and cardiac computed tomography imaging were not performed.

## Conclusion

In this study, patients with severe CT scan lesions were more likely to develop subclinical myocardial damage at mid-term follow-up, despite the absence of biological stigmata of myocardial injury. Early detection of subclinical myocardial dysfunction with speckle tracking echocardiography could be beneficial to ensure early management in recovered patients from COVID-19 infection. Further studies are required to determine the prognostic value of myocardial strain and the long-term evolution of this subclinical myocardial dysfunction in post-COVID-19 patients.

## Data Availability

All data produced in the present study are available upon reasonable request to the authors

## List of abbreviations

CT: computed tomography
IVS: interventricular septum
LVEF: LV ejection fraction
LVEDD: LV end-diastolic diameter
LVEDV: LV end-diastolic volume
LV GLS: left ventricle global longitudinal strain
LVESD: LV end-systolic diameter
LVESV: LV end-systolic volume
RV GLS: right ventricle global longitudinal strain
RT-PCR: real –time reverse –transcription polymerase chain reaction
TAPSE: tricuspid annular plane systolic excursion
TTE: transthoracic echocardiography

